# A cross-sectional study of infection control measures against COVID-19 and psychological distress among Japanese workers

**DOI:** 10.1101/2021.04.16.21255640

**Authors:** Yoshino Yasuda, Tomohiro Ishimaru, Masako Nagata, Seiichiro Tateishi, Hisashi Eguchi, Mayumi Tsuji, Akira Ogami, Shinya Matsuda, Yoshihisa Fujino, for the CORoNaWork project

**Affiliations:** Department of Environmental Epidemiology, Institute of Industrial Ecological Sciences, University of Occupational and Environmental Health, Japan, Kitakyushu, Japan; Department of Occupational Health Practice and Management, Institute of Industrial Ecological Sciences, University of Occupational and Environmental Health, Japan, Kitakyushu, Japan; Department of Occupational Medicine, School of Medicine, University of Occupational and Environmental Health, Japan, Kitakyushu, Japan Department of Psychiatry, University of Occupational and Environmental Health, Japan, Kitakyushu, Japan; Department of Mental Health, Institute of Industrial Ecological Sciences, University of Occupational and Environmental Health, Japan, Kitakyushu, Japan; Department of Environmental Health, School of Medicine, University of Occupational and Environmental Health, Japan, Kitakyushu, Japan; Department of Work Systems and Health, Institute of Industrial Ecological Sciences, University of Occupational and Environmental Health, Japan, Kitakyushu, Japan; Department of Preventive Medicine and Community Health, School of Medicine, University of Occupational and Environmental Health, Japan, Kitakyushu, Japan

**Keywords:** COVID-19, Japan, Occupational Health, Psychological Distress, Prevention and Control

## Abstract

**Objectives:** This study examined the relationship between the status of infection control efforts against COVID-19 in the workplace and workers’ mental health using a large-scale Internet-based study.

**Methods:** This cross-sectional study was based on an Internet monitoring survey conducted during the third wave of the COVID-19 epidemic in Japan. Of the 33,302 people who participated in the survey, 27,036 were included in the analyses. Participants answered whether or not each of 10 different infection control measures were in place at their workplace (e.g. wearing masks at all times during working hours). A Kessler 6 (K6) score of ≥13 was defined as mild psychological distress. The odds ratios (ORs) of psychological distress associated with infection control measures at the workplace were estimated using a multilevel logistic model nested in the prefectures of residence.

**Results:** The OR of subjects working at facilities with 4 or 5 infection control measures for psychological distress was 1.19 (95% confidence interval [CI]: 1.05-1.34, p=0.010), that in facilities with 2 or 3 infection control measures was 1.43 (95% CI: 1.25-1.64, p<0.001), and that in facilities with 1 or no infection control measures was 1.87 (95% CI: 1.63-2.14, p<0.001) compared to subjects whose workplaces had ≥6 infection control measures.

**Conclusion:** Our findings suggest that proactive COVID-19 infection control measures can influence the mental health of workers.

## Introduction

A new type of coronavirus infection (COVID-19) was confirmed in China in December 2019 and spread rapidly around the world. On April 7, 2020, a state of emergency was declared in Tokyo and six other prefectures, followed by a nationwide declaration on April 16. Although the number of infected people temporarily decreased in Japan, a second wave arrived in August of the same year, and a third wave was noted in December to January 2021.

The spread of COVID-19 has transformed people’s lifestyles. To stop the spread of the infection, events and gatherings, including the 2020 Tokyo Olympics, were postponed or cancelled, and restrictions on going out, as well as online schooling and telecommuting were encouraged. The government recommends that residents avoid the “three Cs” of “closed, crowded, and close” in their daily lives by maintaining as much distance between people as possible, going outdoors rather than staying indoors when interacting, and avoiding talking directly to one another whenever possible.^1,2^

In addition to the above measures, various measures are also being taken to prevent infection in the workplace, including maintaining physical distance, wearing masks, basic hygiene (e.g. hand washing), daily health checks, and telework. Further, infection control measures are being implemented not only in public facilities but also in the private sector. Measures to ensure social distance in the workplace, such as spacing out seats, are increasingly common. Even in the hospitality industry, where wearing a mask has historically been frowned upon, the donning of a face mask for health purposes is now routine. Many companies have also introduced programs to allow employees to telecommute, some on a permanent basis. According to a survey,^3^ the number of companies in Tokyo practicing telework has increased markedly, from 24% in March 2020 to 57% in January 2021.

However, with the spread of COVID-19, mental health problems have become an emerging public health issue. Previous studies have reported increased anxiety and mental distress among residents of areas that experienced lockdowns.^4^ Increased rates of mental health problems, such as depression, worsening of existing mental illnesses, suicide, and alcohol dependence, have also been reported.^5–7^ In addition, encouraging avoidance of the three Cs is also suspected to have had a negative impact on the mental health of the public, as such measures necessarily interfere with public interaction, communication, and socialization. Further, anxiety is likely to be heightened by the spread of unconfirmed or even outright false information and the continuous state of fear and worry.^8^ However, while authorities have recommended a variety of measures, the extent of infection prevention measures actually implemented varies among companies. Depending on the industry, some measures may be difficult to implement.

In general, anxiety about infectious diseases such as influenza, HIV, and Middle East Respiratory Syndrome is known to affect mental health,^9–11^ and the same holds true for COVID-19.^12^ A recent review reported that occupational factors and workplace environment were associated with workers’ mental health.^13^ To our knowledge, however, no report has yet examined how the status of infection control efforts against COVID-19 in the workplace influences workers’ mental health.

We hypothesized that the implementation of good infection control efforts in the workplace would have a positive impact on workers’ mental health. Here, we examined the relationship between the status of infection control efforts against COVID-19 in the workplace and workers’ mental health using a large-scale Internet-based study.

## Material and Methods

### Study design and subjects

This cross-sectional study was based on an Internet monitoring survey conducted on December 22nd to 26th, 2020, when the third wave of the COVID-19 epidemic began in Japan. Details of the protocol have already been reported.^14^ In brief, data were collected from workers who had employment contracts at the time of the survey, allocated by prefecture, occupation, and sex. Of the 33,302 people who participated in the survey, 27,036 were included in the study, excluding fraudulent responses. The exclusion criteria were as follows: extremely short response time (≤6 minutes), extremely low body weight (<30 kg), extremely short height (<140 cm), inconsistent answers to similar questions throughout the survey (e.g., inconsistency to questions about marital status and living area), and wrong answers to a staged question used to identify fraudulent responses (choose the third largest number from the following five numbers).

This study was approved by the ethics committee of the University of Occupational and Environmental Health, Japan (reference No. R2-079). Informed consent was obtained via the website.

### Evaluation of infection control measures in the workplace

Participants were asked to answer yes or no concerning whether the following measures had been implemented in their workplace: refraining from and restrictions on business trips; refraining from and restrictions on visitors; refraining from or requesting a limit on the number of people at social gatherings and dinners; refraining from or limiting face-to-face internal meetings; wearing masks at all times during working hours; installing partitions and revising the workplace layout; recommending workers perform daily temperature checks at their homes; encouraging telecommuting; prohibiting workers from eating at their own desk; and requesting employees not come to work when they were not feeling well.

### Assessment of psychological distress

The Kessler 6 (K6) was used to assess psychological distress.^15^ The validity of the Japanese version of the K6 has been confirmed.^16,17^ The K6 was developed to screen for mental disorders, such as depression and anxiety, and is also used as an indicator of the degree of such mental problems, including psychological stress. The K6 is a series of six questions, with scores ranging from 0 (never) to 4 (always), depending on how often the topics of the questions were experienced in the past 30 days. The higher the total score, the greater the possibility of depression or anxiety disorder, with a score of ≥5 indicating the possibility of some kind of depression or anxiety problem and a score of ≥13 indicating a suspected depression or anxiety disorder. In the present study, a K6 score of ≥13 was used as the cut-off value for psychological distress.

### Other covariates

Information on the subject’s socioeconomic status and characteristics of the company, which were considered potential confounding factors, were investigated. Participants noted the following about themselves in an online form: age, sex, prefecture of residence, marital status (married, unmarried, bereaved/divorced), job type (mainly desk work, mainly involving interpersonal communication, and mainly labor), number of employees in the workplace, educational background, equivalent household income (household income divided by the square root of the household size), smoking status, alcohol consumption (6–7 days a week, 4– 5 days a week, 2–3 days a week, less than 1 day a week, hardly ever), telecommuting frequency and subjective evaluation of change in stress and working hours due to COVID-19 (increase, no change, decrease).

### Statistical analyses

The age-sex adjusted and the multivariate adjusted odds ratios (ORs) of psychological distress associated with each infection control measures at the workplace were estimated using a multilevel logistic model nested in the prefecture of residence. The multivariate model was adjusted for sex, age, education, equivalent household income, job type, number of employees at the workplace, smoking status, alcohol consumption, telecommuting frequency, and subjective evaluation of change in stress and working hours due to COVID-19. The incidence rate of COVID-19 during a period between January and December of 2020 by prefecture was also used as a prefecture-level variable. We further estimated the multivariate ORs of psychological distress associated with the number of infection control measures at the workplace.

A p-value of <0.05 was considered statistically significant. All analyses were conducted using Stata (Stata Statistical Software, Release 16; StataCorp LLC, College Station, TX, USA).

## Results

Table 1 shows the basic characteristics according to the number of infection control measures implemented at the workplace. Of the 27036 participants, 14852 (55%) indicated that they had ≥6 infection control measures implemented in their workplace. In contrast, 4177 (15%) of the participants reported having ≤1 infection control measures implemented in their workplace. Compared to the subjects working in companies with fewer infection control measures, the subjects working in companies with more infection control measures tended to be more married, more desk workers, and had higher incomes and education.

**Table 1.** Characteristics of the subjects according to the number of workplace measures against COVID-19

| Factor | Number of workplace measures against COVID-19 (n=27036) |  |  |  |
| --- | --- | --- | --- | --- |
|  | 0-1 | 2-3 | 4-5 | ≥6 |
| Number of subjects | 4177 | 3404 | 4603 | 14852 |
| Age, mean (SD) | 48.6 (9.9) | 47.0 (10.3) | 46.8 (10.6) | 46.6 (10.7) |
| Sex, male | 2489 (59.6%) | 1726 (50.7%) | 2222 (48.3%) | 7377 (49.7%) |
| Marital status, married | 2006 (48.0%) | 1751 (51.4%) | 2462 (53.5%) | 8810 (59.3%) |
| Job type |  |  |  |  |
| Mainly desk work | 1954 (46.8%) | 1475 (43.3%) | 2119 (46.0%) | 7920 (53.3%) |
| Jobs mainly involving interpersonal communication | 870 (20.8%) | 929 (27.3%) | 1342 (29.2%) | 3786 (25.5%) |
| Mainly labor | 1353 (32.4%) | 1000 (29.4%) | 1142 (24.8%) | 3146 (21.2%) |
| Equivalent income (million JPY) |  |  |  |  |
| 50-249 | 1453 (34.8%) | 933 (27.4%) | 990 (21.5%) | 2334 (15.7%) |
| 250-374 | 1197 (28.7%) | 1101 (32.3%) | 1368 (29.7%) | 3884 (26.2%) |
| 375-489 | 838 (20.1%) | 727 (21.4%) | 1116 (24.2%) | 3944 (26.6%) |
| ≥490 | 689 (16.5%) | 643 (18.9%) | 1129 (24.5%) | 4690 (31.6%) |
| Education |  |  |  |  |
| Junior high school | 111 (2.7%) | 83 (2.4%) | 71 (1.5%) | 103 (0.7%) |
| High school | 1475 (35.3%) | 1043 (30.6%) | 1213 (26.4%) | 3222 (21.7%) |
| Vocational school/college, university, graduate school | 2591 (62.0%) | 2278 (66.9%) | 3319 (72.1%) | 11527 (77.6%) |
| Current smoker | 1289 (30.9%) | 990 (29.1%) | 1219 (26.5%) | 3506 (23.6%) |
| Alcohol consumption, 6–7 days a week | 1057 (25.3%) | 755 (22.2%) | 927 (20.1%) | 2935 (19.8%) |
| Number of employees in the workplace |  |  |  |  |
| 1-29 | 2394 (57.3%) | 1248 (36.7%) | 977 (21.2%) | 1546 (10.4%) |
| 30-99 | 1046 (25.0%) | 1203 (35.3%) | 1572 (34.2%) | 3119 (21.0%) |
| 100-999 | 451 (10.8%) | 594 (17.5%) | 1152 (25.0%) | 4956 (33.4%) |
| ≤1000 | 286 (6.8%) | 359 (10.5%) | 902 (19.6%) | 5231 (35.2%) |
| Telecommuting frequency |  |  |  |  |
| More than 4 days per week | 808 (19.3%) | 262 (7.7%) | 259 (5.6%) | 1461 (9.8%) |
| More than 2 days per week | 123 (2.9%) | 108 (3.2%) | 128 (2.8%) | 1118 (7.5%) |
| More than 1 days per week | 63 (1.5%) | 58 (1.7%) | 84 (1.8%) | 673 (4.5%) |
| Less than 1 day per week | 32 (0.8%) | 41 (1.2%) | 81 (1.8%) | 461 (3.1%) |
| Hardly ever | 3151 (75.4%) | 2935 (86.2%) | 4051 (88.0%) | 11139 (75.0%) |
| Change in stress due to COVID-19 epidemic |  |  |  |  |
| increase | 779 (18.6%) | 989 (29.1%) | 1477 (32.1%) | 4834 (32.5%) |
| no change | 3308 (79.2%) | 2332 (68.5%) | 2986 (64.9%) | 9339 (62.9%) |
| decrease | 90 (2.2%) | 83 (2.4%) | 140 (3.0%) | 679 (4.6%) |
| Change in working hours due to COVID-19 epidemic |  |  |  |  |
| increase | 193 (4.6%) | 246 (7.2%) | 372 (8.1%) | 1444 (9.7%) |
| no change | 3654 (87.5%) | 2834 (83.3%) | 3718 (80.8%) | 11725 (78.9%) |
| decrease | 330 (7.9%) | 324 (9.5%) | 513 (11.1%) | 1683 (11.3%) |
| Severe psychological distress (K6≥13) | 464 (11.1%) | 355 (10.4%) | 429 (9.3%) | 1212 (8.2%) |

Table 2 shows the number of infection control efforts put in place and the details of each effort. The most commonly implemented infection control measures were “wearing masks at all times during working hours” (79%) and “refraining from or requesting a limit on the number of people at social gatherings and dinners” (71%). In contrast, relatively few companies had implemented “requesting employees not come to work when they are not feeling well” (9%) and “prohibiting workers from eating at their own desk” (17%).

**Table 2.** Implemented workplace measurements against COVID-19

|  | Number of workplace measures against COVID-19 |  |  |  | Total<br>N=27036 |
| --- | --- | --- | --- | --- | --- |
|  | 0-1 | 2-3 | 4-5 | ≥6 |  |
|  | n=4177 | n=3404 | n=4603 | n=14852 |  |
| Refraining from and restriction of business trips | 56 (1.3%) | 400 (11.8%) | 1479 (32.1%) | 12725 (85.7%) | 14660 (54.2%) |
| Refraining from or restriction on visitors | 26 (0.6%) | 189 (5.6%) | 867 (18.8%) | 11217 (75.5%) | 12299 (45.5%) |
| Refraining from or requesting a limit on the number of people at social gatherings and dinners | 149 (3.6%) | 1200 (35.3%) | 3340 (72.6%) | 14525 (97.8%) | 19214 (71.1%) |
| Refraining from or limiting face-to-face internal meetings | 8 (0.2%) | 250 (7.3%) | 1418 (30.8%) | 13001 (87.5%) | 14677 (54.3%) |
| Wearing masks at all times during working hours | 551 (13.2%) | 2315 (68.0%) | 3970 (86.2%) | 14387 (96.9%) | 21223 (78.5%) |
| Installation of partitions and consideration of workplace layout (e.g. altering desk layout or adjusting flow lines) | 83 (2.0%) | 748 (22.0%) | 2316 (50.3%) | 12587 (84.7%) | 15734 (58.2%) |
| Recommending workers perform daily temperature checks at their homes | 183 (4.4%) | 1349 (39.6%) | 2892 (62.8%) | 12825 (86.4%) | 17249 (63.8%) |
| Encouragement of telecommuting | 114 (2.7%) | 255 (7.5%) | 539 (11.7%) | 6886 (46.4%) | 7794 (28.8%) |
| Prohibiting eating at a worker's own desk | 12 (0.3%) | 87 (2.6%) | 292 (6.3%) | 4206 (28.3%) | 4597 (17.0%) |
| Requesting employees not to come to work when they are not feeling well | 464 (11.1%) | 355 (10.4%) | 429 (9.3%) | 1212 (8.2%) | 2460 (9.1%) |

Table 3 shows the association of the number of infection control measures against COVID-19 and severe psychological distress. As the number of measures increased, the OR for psychological distress decreased. Compared to subjects whose workplaces had implemented 6 or more infection control measures, the OR of subjects with 4 or 5 measures was 1.19 (95% CI: 1.05-1.34, p=0.010), 1.43 (95% CI: 1.25-1.64, p<0.001) for 2 or 3 measures, and 1.87 (95% CI: 1.63-2.14, p<0.001) for 1 or no measures.

**Table 3.**
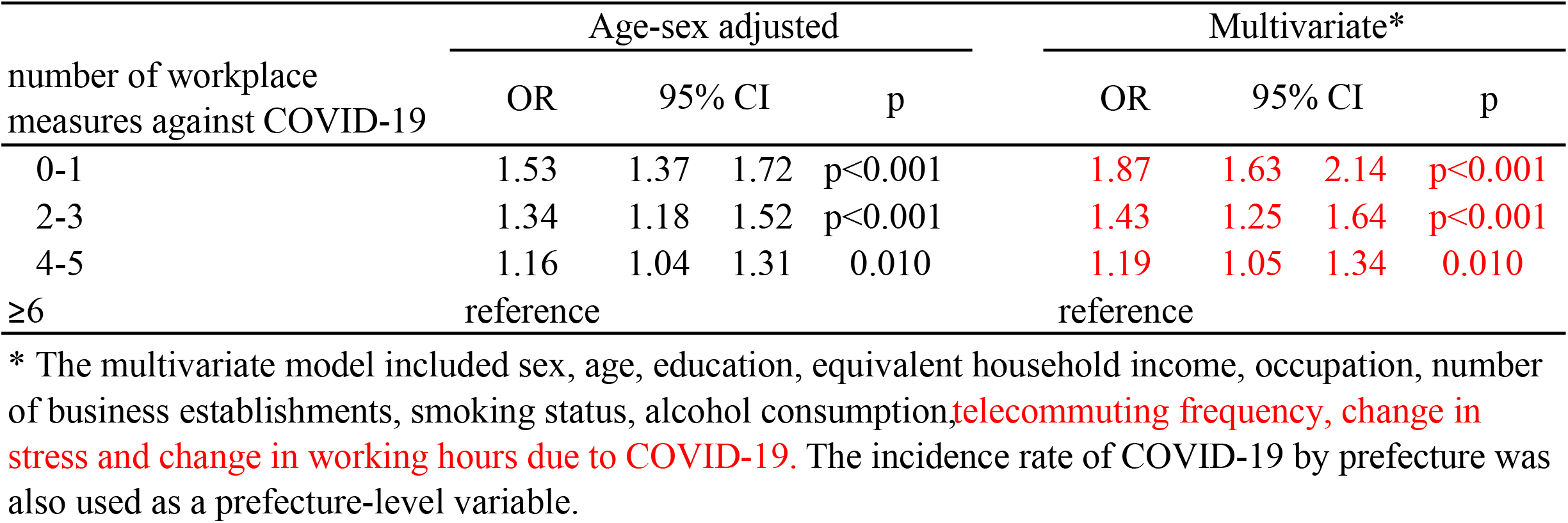
Association of the number of workplace measures against COVID-19 and severe psychological distress (K6≥13)

Table 4 shows the association of each infection control measure and psychological distress. Most of the measures were significantly associated with a reduction in psychological distress, with ORs ranging from approximately 0.70 to 0.80, with statistical significance. In particular, “requesting employees not come to work when they are not feeling well” was associated with a decreased OR for psychological distress (OR=0.56, 95% CI: 0.50-0.62, p<0.001), whereas “prohibiting workers from eating at their own desk” was not associated with psychological distress (OR=1.09, 95% CI:0.97-1.22, p=0.135).

**Table 4.** Association of workplace measures against COVID-19 and severe psychological distresss (K6≥13)

|  | Age-sex adjusted |  |  |  | Multivariate* |  |  |  |
| --- | --- | --- | --- | --- | --- | --- | --- | --- |
|  | OR | 95% CI |  | p | OR | 95% CI |  | p |
| Refraining from and restrictions on business trips | 0.78 | 0.72 | 0.85 | <0.001 | 0.75 | 0.68 | 0.83 | <0.001 |
| Refraining from and restrictions on visitors | 0.85 | 0.78 | 0.92 | <0.001 | 0.82 | 0.75 | 0.90 | <0.001 |
| Refraining from or requesting a limit on the number of people at social gatherings and dinners | 0.75 | 0.68 | 0.81 | <0.001 | 0.70 | 0.64 | 0.78 | <0.001 |
| Refraining from or limiting face-to-face internal meetings | 0.77 | 0.70 | 0.83 | <0.001 | 0.74 | 0.67 | 0.81 | <0.001 |
| Wearing masks at all times during working hours | 0.82 | 0.74 | 0.91 | <0.001 | 0.74 | 0.66 | 0.83 | <0.001 |
| Installation of partitions and consideration of workplace layout (e.g. altering desk layout or adjusting flow lines) | 0.79 | 0.72 | 0.86 | <0.001 | 0.75 | 0.68 | 0.82 | <0.001 |
| Recommending workers perform daily temperature checks at their homes | 0.82 | 0.75 | 0.90 | <0.001 | 0.77 | 0.70 | 0.85 | <0.001 |
| Encouragement of telecommuting | 0.85 | 0.77 | 0.93 | 0.001 | 0.85 | 0.76 | 0.96 | 0.009 |
| Prohibiting workers from eating at their own desk | 1.08 | 0.97 | 1.20 | 0.172 | 1.09 | 0.97 | 1.22 | 0.135 |
| Requesting employees not come to work when they are not feeling well | 0.60 | 0.55 | 0.65 | <0.001 | 0.56 | 0.50 | 0.62 | <0.001 |

\* The multivariate model included sex, age, education, equivalent household income, occupation, number of business establishments, smoking status, alcohol consumption, telecommuting frequency, change in stress and change in working hours due to COVID-19. The incidence rate of COVID-19 by prefecture was also used as a prefecture-level variable.

## Discussion

This study demonstrated an association between psychological distress and the implementation of 10 common COVID-19 infection prevention measures recommended in the workplace. In addition, our study revealed that the more infection control measures implemented in the workplace, the lower the workers’ mental distress.

Each individual preventive measure was found to be associated with low psychological distress. In particular, the policy of “requesting employees not come to work when they are not feeling well” markedly reduced psychological distress. This measure, which likely reduces the risk of infection of others when one employee is infected, was generally thought by workers to be directly effective in minimizing damage in the event of infection around them, and was linked to their mental stability. Sickness presenteeism is reportedly influenced by organizational culture, employment instability, and socioeconomic status.^18–20^ Such a clear statement of company policy would discourage workers with a poor socioeconomic status against concealing their fever and coming to work, thereby preventing infection.

In contrast, only the measure “prohibiting workers from eating at their own desk” was not associated with decreased psychological distress, possibly reflecting the psychological distress caused by reducing communication with peers during a time of refreshment. These results suggest that communication is becoming an issue during the COVID-19 pandemic and that efforts to consciously balance COVID-19 infection control and communication in the workplace are needed.^21^ It should be noted, however, that interpretation of these results needs to be caution. These analyses compared people who took a certain measure with those who did not, but the people who did not take such measure may also take other measures. Therefore, it does not evaluate the effectiveness of a particular measure in isolation.

The present study revealed marked variation in the COVID-19 infection control measures among companies. These differences in among companies are attributed to various factors, including the difficulty of adopting some measures in certain industries, the influence of the sense of risk among companies and employers, and the costs associated with the implementation of measures. This is supported by the results of this study. Those who worked in companies that implemented more infection control measures had higher incomes, more education, and larger company sizes. The feasibility of infection control measures also differs depending on the work environment. For example, in workplaces that do not have break rooms for taking meals, prohibiting workers from eating at their own desks may be difficult. In workplaces where employees have fixed starting and working hours, crowding of changing rooms at certain times may be unavoidable. The ability to implement such measures varies depending on the budget available, the size of the company, and the presence of an individual in charge of promoting safety and health.

Our study also found that the more infection control measures implemented in the workplace, the less substantial workers’ mental distress. We assumed that companies with more infection control measures in place would have stronger company governance. Such proactive corporate initiatives for infection control may contribute to the mental health of workers by promoting a safety climate and psychosocial safety climate in the workplace.^22^ The results suggest that the implementation of proactive infection control helps reduce psychological distress. Infection anxiety concerning COVID-19 reportedly affects mental health,^5,7,8,12,23^ which is consistent with findings for other infectious diseases, such as hepatitis and HIV.^24,25^ These results suggest that infection control measures alleviate workers’ anxiety about infection. Proactive infection prevention measures in the workplace are expected to reduce workers’ anxiety about infection and thereby have a positive impact on workers’ mental health.^26^

Several possible reasons may explain why active infection control in the workplace is associated with psychological distress in workers. As mentioned above, infection prevention measures have a direct effect on reducing infection anxiety. In addition, the implementation of appropriate COVID-19 control measures based on correct information can itself serve as a form of infection prevention education for workers.^27^ When an organization faces a crisis, whether or not the employer provides appropriate support affects the mental health of the workers.^23^ A clear COVID-19 infection prevention policy in the workplace can build a relationship of trust between the employer and workers and consequently reduce workers’ psychological distress.^13^

Several limitations associated with the present study warrant mention. First, this study was a cross-sectional study, so causality is unclear. More psychologically anxious people may have been prone to underestimate the efficacy of the infection control measures in the workplace. However, since this study inquired about the existence of visible and physical measures, we believe that the likelihood of wrong answers was low. Second, we did not confirm the time when the countermeasures were implemented. This study investigated the situation as of December 2020, when the infection was at its peak in Japan. Therefore, the degree of the subjects’ anxiety and their reaction to their workplace’s efforts may have differed between the time when the infection rate was relatively low and the time of the survey, when the rate was relatively high. Third, a lack of information regarding possible confounding factors, such as industry and employment stability might affect the results. For example, we did not ask about the subjects’ workplaces or industries in detail, but some of these factors likely affected the results, such as the size of the workplace space (small office vs. a large space, such as a retail store) or how often the subjects were in contact with unspecified numbers of people. Finally, establishments that have implemented many measures are more likely to be companies that are enthusiastic about mental health measures and consider their employees’ well-being on a regular basis, so the mental distress may have already been low at baseline; however, this aspect was not evaluated.

This study suggests that proactive COVID-19 infection control measures can lead to improved mental health care for workers. Proactive infection prevention measures in the workplace are expected to reduce workers’ anxiety about infection and have a positive impact on workers’ mental health. The implementation of COVID-19 infection control measures in the workplace is recommended not only as a way to prevent infection but also as a new mental health measure.

## Data Availability

Data not available due to ethical restrictions

## Acknowledgements

This study was supported and partly funded by the University of Occupational and Environmental Health, Japan; General Incorporated Foundation (Anshin Zaidan); The Development of Educational Materials on Mental Health Measures for Managers at Small-sized Enterprises; Health, Labour and Welfare Sciences Research Grants; Comprehensive Research for Women’s Healthcare (H30-josei-ippan-002); Research for the Establishment of an Occupational Health System in Times of Disaster (H30-roudou-ippan-007), scholarship donations from Chugai Pharmaceutical Co., Ltd., the Collabo-Health Study Group, and Hitachi Systems, Ltd.

The current members of the CORoNaWork Project, in alphabetical order, are as follows: Dr. Yoshihisa Fujino (present chairperson of the study group), Dr. Akira Ogami, Dr. Arisa Harada, Dr. Ayako Hino, Dr. Hajime Ando, Dr. Hisashi Eguchi, Dr. Kazunori Ikegami, Dr. Kei Tokutsu, Dr. Keiji Muramatsu, Dr. Koji Mori, Dr. Kosuke Mafune, Dr. Kyoko Kitagawa, Dr. Masako Nagata, Dr. Mayumi Tsuji, Ms. Ning Liu, Dr. Rie Tanaka, Dr. Ryutaro Matsugaki, Dr. Seiichiro Tateishi, Dr. Shinya Matsuda, Dr. Tomohiro Ishimaru, and Dr. Tomohisa Nagata. All members are affiliated with the University of Occupational and Environmental Health, Japan.

## Disclosure

### Ethical approval

This study was approved by the ethics committee of the University of Occupational and Environmental Health, Japan (reference No. R2-079).

### Informed Consent

Informed consent was obtained through an online website.

### Registry and the Registration No. of the study/Trial

N/A

### Animal Studies

N/A

